# Mendelian randomization analysis identified genes pleiotropically associated with the risk and prognosis of COVID-19

**DOI:** 10.1101/2020.09.02.20187179

**Authors:** Di Liu, Jingyun Yang, Bowen Feng, Wenjin Lu, Chuntao Zhao, Lizhuo Li

## Abstract

**Objectives:** COVID-19 has caused a large global pandemic. Patients with COVID-19 exhibited considerable variation in disease behavior. Pervious genome-wide association studies have identified potential genetic variants involved in the risk and prognosis of COVID-19, but the underlying biological interpretation remains largely unclear.

**Methods:** We applied the summary data-based Mendelian randomization (SMR) method to identify genes that were pleiotropically associated with the risk and various outcomes of COVID-19, including severe respiratory confirmed COVID-19 and hospitalized COVID-19.

**Results:** In blood, we identified 2 probes, ILMN_1765146 and ILMN_1791057 tagging *IFNAR2*, that showed pleiotropic association with hospitalized COVID-19 (β [SE] = 0.42 [0.09], *P* = 4.75×10^−06^ and β [SE] = –0.48 [0.11], *P* = 6.76×10^−06^, respectively). Although no other probes were significant after correction for multiple testing in both blood and lung, multiple genes as tagged by the top 5 probes were involved in inflammation or antiviral immunity, and several other tagged genes, such as *PON2* and *HPS5*, were involved in blood coagulation.

**Conclusions:** We identified *IFNAR2* and other potential genes that could be involved in the susceptibility or prognosis of COVID-19. These findings provide important leads to a better understanding of the mechanisms of cytokine storm and venous thromboembolism in COVID-19 and potential therapeutic targets for the effective treatment of COVID-19.

## Introduction

Coronavirus disease 2019 (COVID-19), which is caused by the severe acute respiratory syndrome coronavirus 2 (SARS-CoV-2), has created a large global pandemic and poses a serious threat to public health (1, 2). As of August 7, 2020, there were more than 19.3 million confirmed cases worldwide, with the total deaths exceeding 719,830 (3). SARS-CoV-2 is a highly pathogenic and transmissible coronavirus that primarily spreads through respiratory droplets and close contact (4). Seeking solutions to control the spread of COVID-19 and exploring effective treatments are of utmost importance to address the global challenge posed by COVID-19. Therefore, there is pressing urgency to further identify the pathological mechanisms underlying COVID-19.

Patients with COVID-19 exhibited considerable variation in disease behavior. Recently, genome-wide association studies (GWAS) have been performed to identify genetic variants associated with diagnosis of and prognosis of COVID-19 (5, 6), but biological interpretation of their findings remains largely unclear. Previous research found that approximately 88% of trait-associated genetic variants detected by GWAS resided in non-coding regions of the genome and might have regulatory functions on gene expression (7). In the context of COVID-19 research, it is therefore important to explore genes whose expressions were pleiotropically/potentially causally associated with susceptibility or the development of COVID-19.

Different from conventional randomized controlled trials (RCTs), Mendelian randomization (MR), which uses genetic variants as the proxy to randomization (8), is a promising tool to search for pleotropic/potentially causal effect of an exposure (e.g., gene expression) on the outcome (e.g., COVID-19 susceptibility). MR minimizes confounding and reverse causation that are commonly encountered in traditional association studies (8, 9), and has been successful in identifying gene expression sites or DNA methylation loci that are pleiotropically/potentially causally associated with various phenotypes, such as cardiovascular diseases, BMI, and rheumatoid arthritis (10–13).

In this paper, we applied the summary data-based MR (SMR) method integrating summarized GWAS data for COVID-19 and cis- eQTL (expression quantitative trait loci) data to prioritize genes that are pleiotropically/potentially causally associated with the risk and prognosis of COVID-19.

## Methods

### Data sources

#### eQTL data

In the SMR analysis, cis-eQTL genetic variants were used as the instrumental variables (IVs) for gene expression. We performed SMR analysis for gene expression in blood and lung separately. For blood, we used the CAGE eQTL summarized data (14), which included 2765 participants. For lung, we used the V7 release of the GTEx eQTL summarized data (15), which included 278 participants. The eQTL data for blood and lung can be downloaded at https://cnsgenomics.com/data/SMR/#eQTLsummarydata.

#### GWAS data for COVID-19

The GWAS summarized data were provided by *the COVID-19 host genetics initiative* (6) and can be downloaded at https://www.covid19hg.org/results/. Three phenotypes were examined, including severe respiratory confirmed COVID-19, COVID-19 and hospitalized COVID-19. Details on definition of the phenotypes can be found in **Table S1**. The control groups were subjects from the general population without the specific phenotype, subjects who were COVID-19 negative based on prediction or self-report, or subjects who had COVID-19 without hospitalization, making a total of five comparisons: severe respiratory confirmed COVID-19 (n = 536) vs. population (n = 329,391; hereafter severe COVID-19); COVID-19 (n = 6,696) vs. population (n = 1073072; hereafter COVID-19), COVID-19 (n = 3,523) vs. COVID-19 negative (n = 36,634; hereafter COVID-19 negative); hospitalized COVID-19 (n = 3,199) vs. population (n = 897,488; hereafter hospitalized COVID-19), and hospitalized COVID-19 (n = 928) vs. COVID-19 without hospitalization (n = 2,028; hereafter COVID-19 without hospitalization). In addition, we repeated the SMR analysis for severe respiratory confirmed COVID-19 using GWAS summarized data from the Severe COVID-19 GWAS Group (5) (hereafter severe COVID-19 NEJM). The study included 1,160 patients who had severe respiratory confirmed COVID-19, and 2,205 participants from the general population without COVID-19 as the control (**Table 1**). The definition of severe respiratory confirmed COVID-19 in this study was different from the one as defined by *the COVID-19 host genetics initiative* (**Table S1**). The GWAS summarized data can be downloaded at https://ikmb.shinyapps.io/COVID-19_GWAS_Browser/.

**Table 1.**
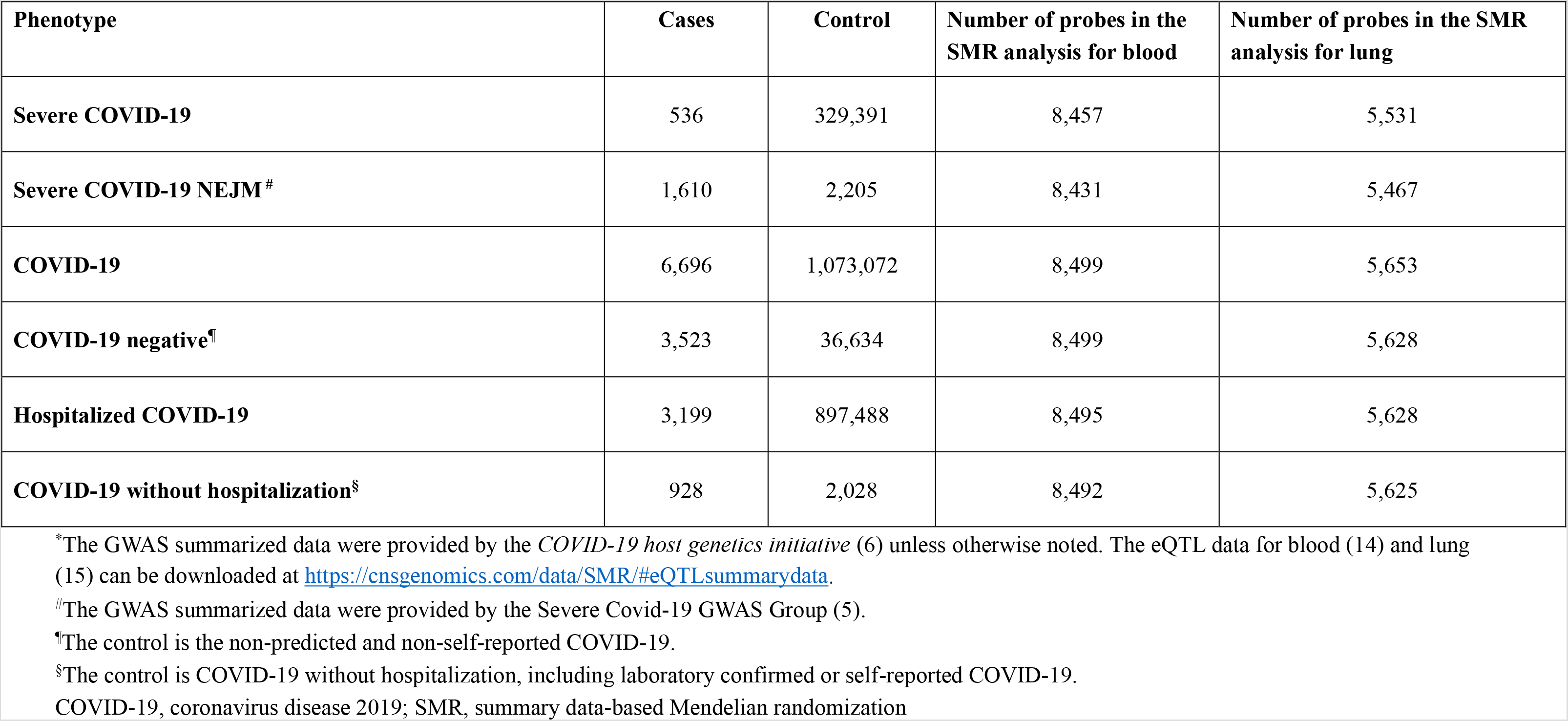
Basic information of the GWAS data and eQTL data in the SMR analyses*.

## SMR analysis

MR was undertaken with cis-eQTL as the IV, gene expression as the exposure, and each of the three phenotypes as the outcome (in comparison with different control groups). MR analysis was done using the method as implemented in the software SMR. Detailed information regarding the SMR method was reported in a previous publication (10). In brief, SMR applies the principles of MR to jointly analyze GWAS and eQTL summary statistics in order to test for pleotropic association between gene expression and a trait due to a shared and potentially causal variant at a locus. The heterogeneity in dependent instruments (HEIDI) test was performed to evaluate the existence of linkage in the observed association. Rejection of the null hypothesis (i.e., *P*_HEIDI_< 0.05) indicates that the observed association might be due to two distinct genetic variants in high linkage disequilibrium with each other. We adopted the default settings in SMR (e.g., *P*_eQTL_ < 5 × 10^−8^, minor allele frequency [MAF] > 0.01, removing SNPs in very strong linkage disequilibrium [LD, r^2^> 0.9] with the top associated eQTL, and removing SNPs in low LD or not in LD [r^2^< 0.05] with the top associated eQTL), and used false discovery rate (FDR) to adjust for multiple testing.

Annotations of transcripts were based on the Affymetrix exon array S1.0 platforms. To functionally annotate putative transcripts, we conducted functional enrichment analysis using the functional annotation tool “Metascape” (16) for the top tagged genes in blood and lung, separately. Gene symbols corresponding to putative genes *(P*< 0.05) were used as the input of the gene ontology (GO) and Kyoto Encyclopedia of Genes and Genomes (KEGG) enrichment analysis.

Data cleaning and statistical/bioinformatical analysis was performed using R version 4.0.0 (https://www.r-project.org/), PLINK 1.9 (https://www.coggenomics.org/plink/1.9/) and SMR ((https://cnsgenomics.com/software/smr/).

## Results

### Basic information of the summarized data

The number of cases and controls varied dramatically among different analyses. The number of probes were approximately 8,500 and 5,500 in the analysis for blood and lung, respectively. The detailed information was shown in **Table 1**.

### SMR analysis in blood

Information of the top 5 probes for each phenotype was presented in **Table 2**. Using GWAS summarized data from *the COVID-19 host genetics initiative*, we identified 2 probes, ILMN_1765146 and ILMN_1791057 tagging *IFNAR2*, that showed pleiotropic association with hospitalized COVID-19 (β [SE] = 0.42 [0.09], *P* = 4.75×10^−06^ and β [SE] = –0.48 [0.11], *P* = 6.76×10^−06^, respectively), but no significant pleiotropic association for other phenotypes after correction for multiple testing **(Figure 1, Figure S1–5)**. The same two probes were also among the top 5 probes in the analysis of severe COVID-19 NEJM and COVID-19, although they did not reach statistical significance after correction for multiple testing (**Table 2**). In addition, we found that multiple probes tagging *TRIM5* were among the top 5 probes in the analysis of severe COVID-19 NEJM and hospitalized COVID-19 in blood.

**Figure 1.**
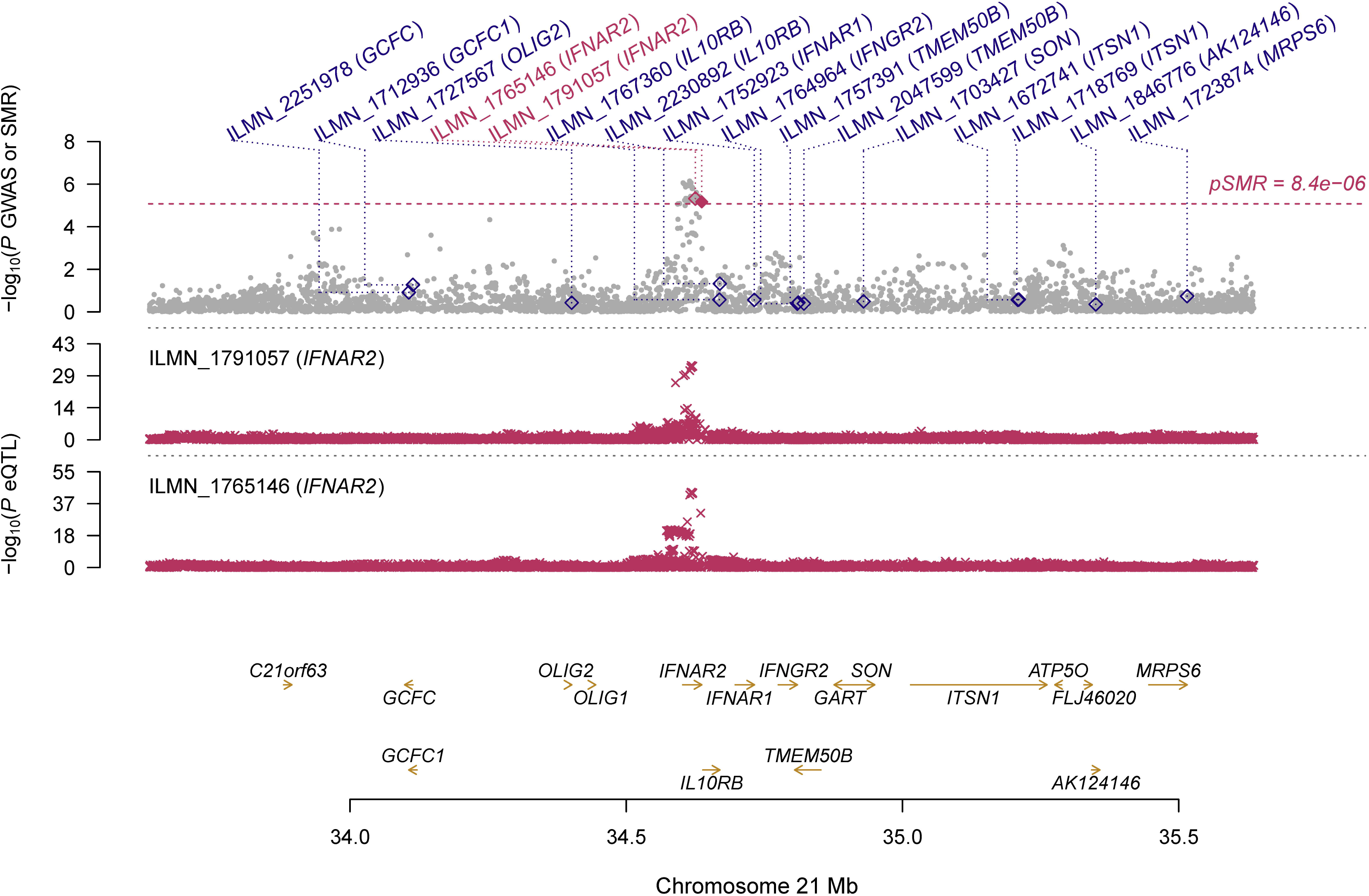
Prioritizing genes around *IFNAR2* in association with hospitalized COVID-19 in blood. Top plot, grey dots represent the –log_10_*(P* values) for SNPs from the GWAS of hospitalized COVID-19, and rhombuses represent the –log_10_*(P* values) for probes from the SMR test with solid rhombuses indicating that the probes pass HEIDI test and hollow rhombuses indicating that the probes do not pass the HEIDI test. Middle plot, eQTL results in blood for two probes, ILMN_1765146 and ILMN_1791057 probes, tagging *IFNAR2*. Bottom plot, location of genes tagged by the probes. Highlighted in maroon indicates probes that pass SMR threshold. COVID-19, coronavirus disease 2019; *IFNAR2*, interferon alpha and beta receptor subunit 2; GWAS, genome-wide association studies; SMR, summary data-based Mendelian randomization; HEIDI, heterogeneity in dependent instruments; eQTL, expression quantitative trait loci

**Table 2.**
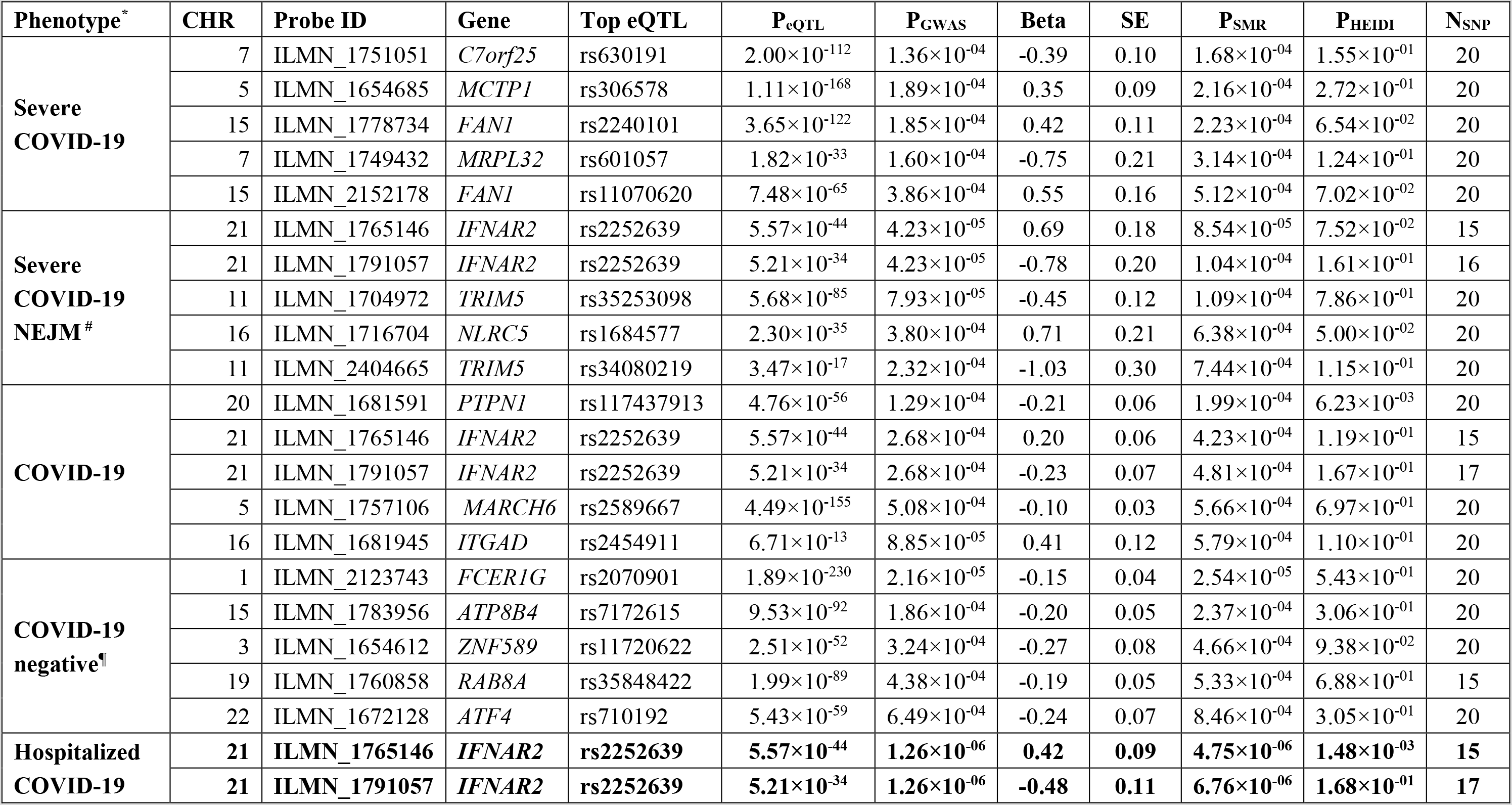
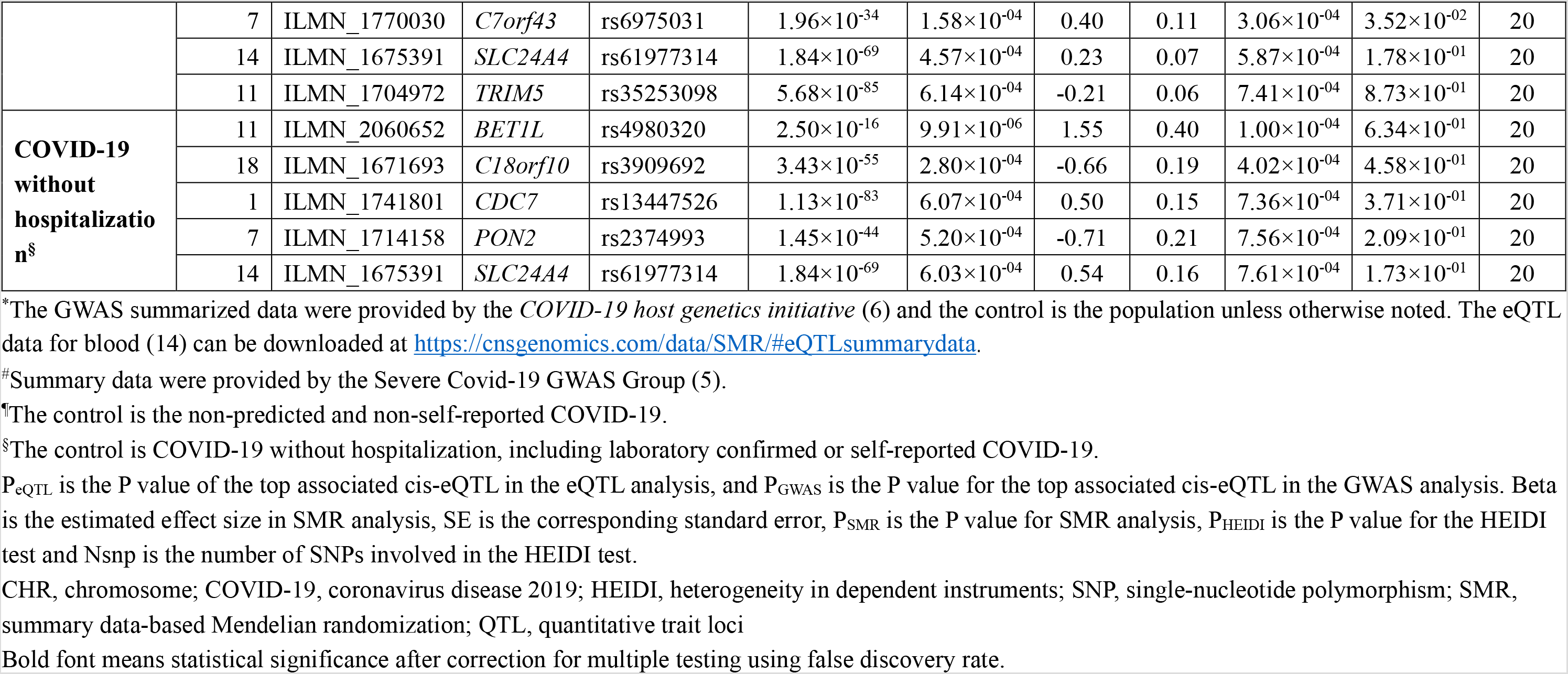
The top five probes identified in the SMR analysis for blood.

GO enrichment analysis of biological process and molecular function showed that the genes tagged by the top 5 probes were involved in four GO terms, including regulation of type I interferon-mediated signaling pathway (GO: 0060338), regulation of vesicle-mediated transport (GO: 0060627), response to endoplasmic reticulum stress (GO: 0034976); and response to oxidative stress (GO:0006979). Multiple genes as tagged by the top 5 probes, including *IFNAR2, TRIM5, NLRC5, MCTP1, PTPN1, FCER1G, ATF4* and *PON2*, were involved in inflammation or antiviral immunity (**Table S2**).

### SMR analysis in lung

We didn’t identify any significant pleiotropic association after correction for multiple testing **(Figure S6–11)**. Information of the top 5 probes for each phenotype was presented in **Table 3**. We found that 3 probes tagging *AP006621* were among the top probes in the analysis of severe COVID-19 NEJM (**Table 3**). In addition, multiple genes, including *C7orf25, ITGAD, FCER1G* and *ZNF589*, were tagged by the top 5 probes in both blood and lung.

**Table 3.**
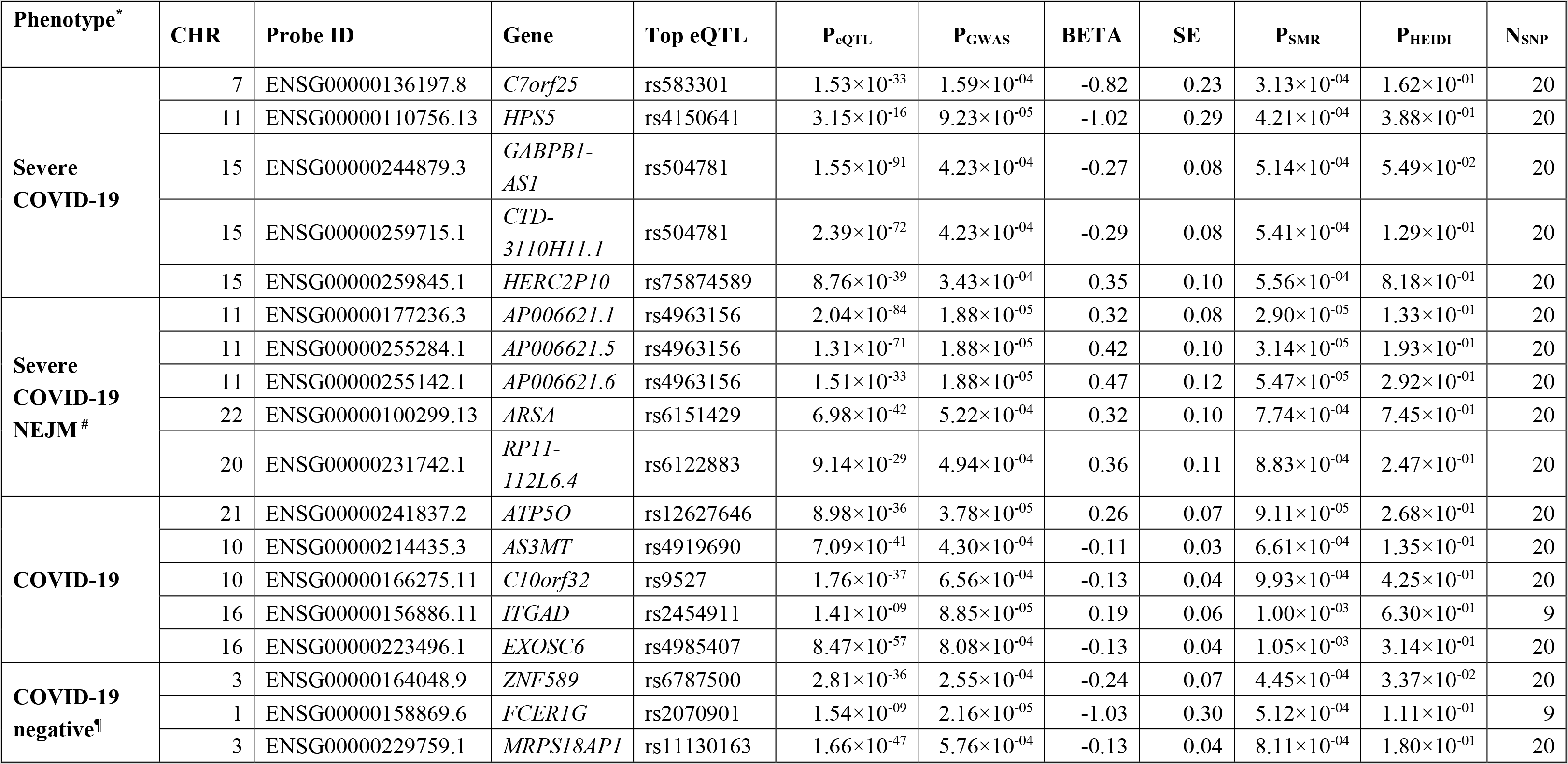
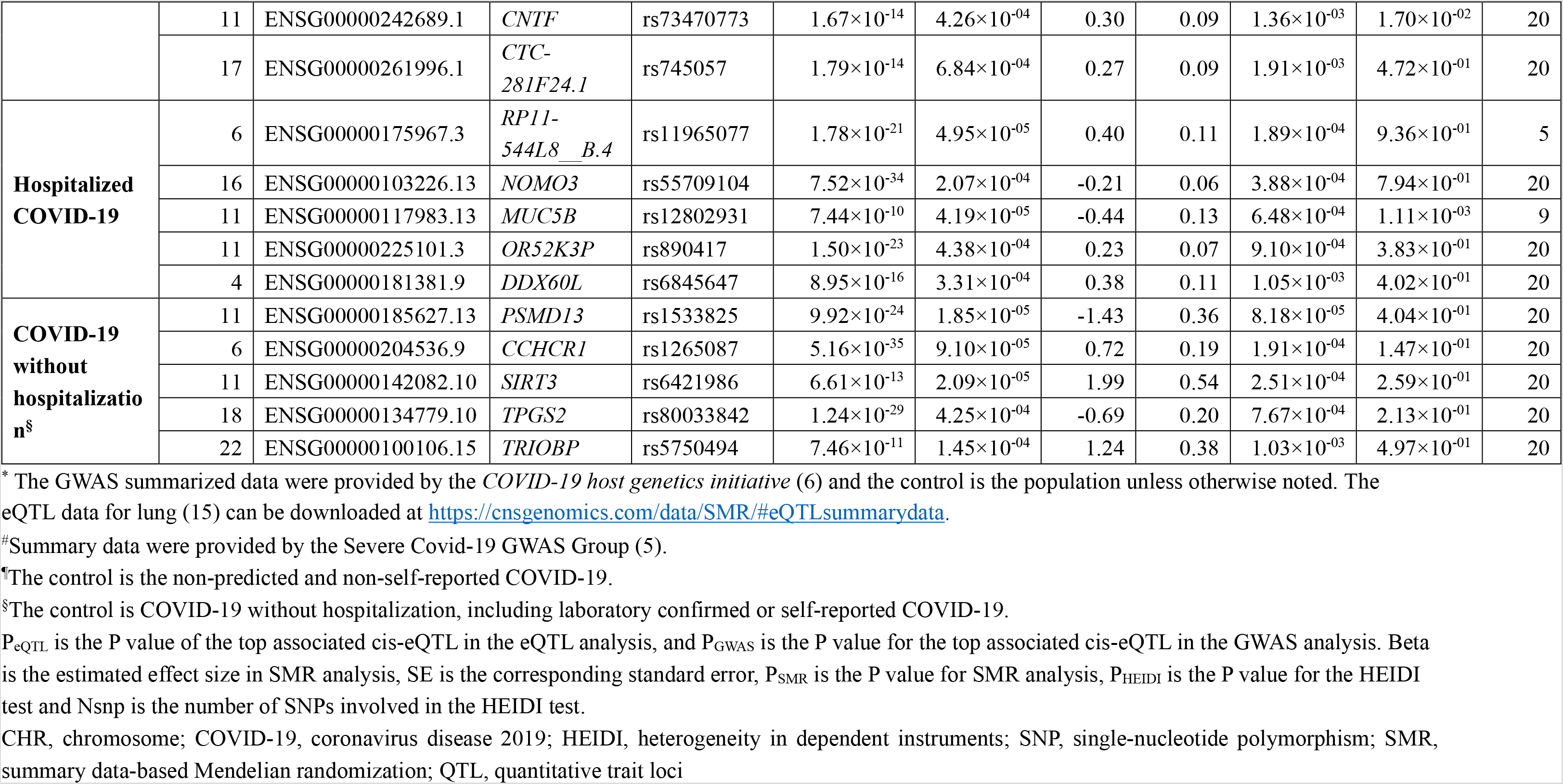
The top five probes identified in the SMR analysis for lung.

The genes tagged by the top 5 probes were involved in two GO terms, including stimulatory C-type lectin receptor signaling pathway (GO: 0002223) and leukocyte activation involved in immune response (GO: 0002366). Multiple genes as tagged by the top 5 probes, including *ARSA, FCER1G, XOSC6* and *PSMD13*, were involved inflammation or antiviral immunity (**Table S3**).

## Discussion

In the present study, we integrated GWAS and eQTL data in the MR analysis to explore putative genes that showed pleiotropic/potentially causal association the susceptibility/prognosis of COVID-19. We identified 2 probes tagging *IFNAR2* showing pleiotropic association with hospitalized COVID-19 in blood. Multiple genes as tagged by the top 5 probes were involved in inflammation and antiviral immunity in both blood and lung. Several genes tagged by the top probes were in blood coagulation. Our findings provided important leads to a better understanding of the mechanism of cytokine storm and venous thromboembolism in COVID-19 and revealed potential therapeutic targets for the effective treatment of COVID-19.

Interferons (IFNs) refer to a group of signaling proteins made and released by the host cells in response to viral invasion (17). There are three types of IFNs: type I IFNs (IFN-α/β), type II IFNs (IFN-γ) and type III IFNs (IFN-λ) (18–20). *IFNAR2* (interferon alpha and beta receptor subunit 2), located at 21q22.11, encodes one of the two type I IFNs (21). In the cascade of host’s response to coronavirus, IFNs play an essential role in the establishment of antiviral state and in intensifying the antiviral response (22). It was found that IFNs could have both beneficial and detrimental effect on SARS-Cov-2 replication (23). A recent retrospective study of 77 adults found that IFN-α2b treatment with or without arbidol significantly reduced the duration of detectable virus in the upper respiratory tract of COVID-19 patients (24). However, the COVID-19 Treatment Guidelines Panel recommend against the use of interferons for the treatment of patients with severe and critical COVID-19, except in a clinical trial, due to insufficient data to support the beneficial or detrimental effects of interferons (25). Because *IFNAR2* was tagged by the top 5 probes for multiple phenotypes, we think that it is likely involved in determining COVID-19 severity and could be a potential therapeutic target for the treatment of COVID-19. More studies are needed to elucidate the mechanisms underlying the dual role of IFNs during SARS-Cov-2 and whether/how *IFNAR2* is involved in this process.

We found that multiple genes as tagged by the top 5 probes were involved in inflammation. *ATF4* (activating transcription factor 4) is an endoplasmic reticulum stress sensor that defends lungs via induction of heme oxygenase 1 (26). ATF4 were decreased in inflamed intestinal mucosa from patients with active Crohn’s disease or active ulcerative colitis, and ATF4 deficiency promotes intestinal inflammation in mice (27). *ATF4* was downregulated in the alveolar type II cells of the elderly, compared with the young (28). *ATF4* has more than 20 candidate downstream factors, with the majority of them being significantly downregulated in the elderly, who demonstrated compromised ATF4-dependent ability to respond to endoplasmic reticulum stress. Lung-specific delivery of ATF4-related antioxidants has the potential to work in synergy with promising antiviral drugs to further improve COVID-19 outcomes in the elderly (28). Some of the tagged genes, such as *TRIM5, NLRC5, MCTP1, PTPN1, ARSA* and *FCER1G*, are also involved antiviral immunity (29–35). These genes have not been reported to be associated with COVID-19 in previous studies.

It was estimated that approximately 40% of COVID-19 patients were considered as having a high risk of venous thromboembolism risk, and 11% of them developed venous thromboembolism without prophylaxis (36). Abnormal coagulation in patients with COVID-19 was associated with an increased risk of death (37). Recent research indicated that dysregulated platelets and neutrophils cooperated to drive a systemic prothrombotic state in SARS-CoV-2 infection (38). However, the underlying mechanisms in general, and genetic contributions in particular, remain to be further explored. In our study, we found two genes, as tagged by the top five probes, that were involved in blood coagulation: *PON2* and *HPS5* in the analysis of COVID-19 without hospitalization in blood (**Table 2**) and severe COVID-19 in lung (**Table 3**), respectively. *PON2* (paraoxonase 2) encodes a member of the paraoxonase gene family, and acts as a cellular antioxidant, protecting cells from oxidative stress (39). It was shown that deregulated redox regulation in PON2 deficiency caused vascular inflammation and abnormalities in blood coagulation (40). *HPS5* encodes a protein that is involved in organelle biogenesis associated with melanosomes and platelet dense granules (41). Mutations in *HPS5* are associated with Hermansky-Pudlak syndrome type 5 (HPS5), a subtype of a series of disorders characterized by oculocutaneous albinism and prolonged bleeding (42). Although the pleotropic association was not statistically significant in the SMR analysis after correction for multiple testing (P = 7.56×10^−4^ for *PON2* and 4.21×10^−4^ for *HPS5)*, our findings suggested potential involvement of these genes in the pathogenesis of venous thromboembolism in COVID-19 patients. More studies are needed to explore the functions of these two genes in response to SARS-CoV-2 infection.

Our study has some limitations. The GWAS analyses did not control confounding factors which might affect the outcome. It is also unclear whether selection of the subjects in the GWAS studies was a representative of the exposure-outcome distributions in the overall population, and therefore, the possibility of selection bias, which can affect estimation accuracy, could not be ruled out. The GWAS studies only examined the short-term effect of COVID-19 due to the limited duration of the COVID-19 pandemic, and we were unable to assess the long-term outcomes/lingering effects of COVID-19. Similarly, we could not analyze the genetic contribution of other interesting phenotypes, such as different disease behaviors among children/teens, adults and the elderly patients, and asymptomatic COVID-19, due to a lack of the corresponding GWAS summarized data. We only performed analyses using blood and lung eQTL data, more studies are needed to explore tissue-and cell-type-specific genes involved in the host responses to COVID-19 infection. Due to a lack of individual eQTL data, we could not quantify the changes in gene expression in patients with COVID-19 in comparison with the control.

## Conclusion

We identified *IFNAR2* and other potential genes that could be involved in the susceptibility/prognosis of COVID-19. These findings provide important leads to a better understanding of the mechanism of cytokine storm in COVID-19 and reveals potential therapeutic targets for the effective treatment of COVID-19.

## Data Availability

All data generated or analyzed during this study are included in this published article and its supplementary information files.

## Acknowledgements

Dr. Lizhuo Li’s research was supported by the National Natural Science Foundation of China (Grant No. 81660329). The study was supported by NIH/NIA grants P30AG10161, R01AG15819, R01AG17917, R01AG36042, U01AG61356 and 1RF1AG064312–01. Di Liu was supported by China Scholarship Council (CSC 201908110339).

The authors confirmed that all authors have reviewed the contents of the article being submitted, approved its contents, and validated the accuracy of the data.

## Author contribution

DL, JY and LL designed and registered the study. DL, BF and WL analyzed data and performed data interpretation. DL and JY wrote the initial draft and BF, WL, CZ and LL contributed writing to subsequent versions of the manuscript. All authors reviewed the study findings and read and approved the final version before submission.

